# The combination of *SPP1* knockdown and gemcitabine treatment enhances apoptosis and reduces invasiveness of pancreatic cancer cells

**DOI:** 10.1101/2024.05.05.24306611

**Authors:** Ntombikayise Xelwa, Previn Naicker, Jones Omoshoro-Jones, John Devar, Martin Smith, Geoffrey Candy, Tanya Nadine Augustine, Ekene Emmanuel Nweke

## Abstract

**Background:** Pancreatic ductal adenocarcinoma (PDAC) is poised to be a leading cause of cancer-related deaths. Despite developing new treatment strategies, patient outcomes have not significantly improved. Chemoresistance has been implicated as a major contributor to ineffective treatments observed with studies suggesting combination therapy targeting multiple pathways. This study explored dysregulated genes in tumours of PDAC patients to identify targets which could be used effectively in combination with conventional therapy against cancer cells.

**Methods:** In this study, PCR arrays were used for gene expression profiling of tumours obtained from South African PDAC patients to identify key differentially expressed pathways and potentially new therapeutic target genes. *SPP1* was selected and RNA interference was used to knock the gene down. Migration and apoptosis assays were used to evaluate the effect of the knockdown, alone and in combination with gemcitabine, on a pancreatic cancer cell line, MIA PaCa-2. Proteomic analysis using SWATH-MS was used to demonstrate potential molecular mechanisms linked to the morphological and phenotypical effects observed with treatment.

**Results:** We demonstrated several genes linked to the growth factor and signal transduction signalling pathways, and identified *SPP1* as a target. We observed that by combining *SPP1* knockdown with conventional chemotherapy, gemcitabine, resulted in a synergistic effect, leading to an enhanced early apoptotic response. A decline of migratory and invasive capabilities of MIA PaCa-2 cells was observed upon subjecting the cancer cells to *SPP1* reduction and gemcitabine treatment. Furthermore, proteomic analyses uncovered several pathways that were dysregulated by the combination therapy including both pro-and anti-tumorigenic ones.

**Conclusion:** The study findings indicate that *SPP1* could be a potential therapeutic target for PDAC, and the possible synergistic effects observed when *SPP1* knockdown was combined with gemcitabine treatment suggest a potential avenue for developing more effective treatments for PDAC while exploring tumour cell adaptation for survival.

## Introduction

Pancreatic ductal adenocarcinoma (PDAC) represents almost 90% of all pancreatic cancers (Skorupan *et al*., 2023). Pancreatic cancer is among the top three leading causes of cancer-related deaths in both sexes in the USA (Siegel *et al*., 2023), with almost the same number of new cases and deaths reported annually.

Treatment strategies for PDAC typically include surgery, chemotherapy or radiotherapy. Resistance to chemotherapy is one of the major challenges in treating PDAC (Jadid *et al*., 2021; Heiat *et al*., 2023). Chemotherapeutic drugs used in the treatment of PDAC include gemcitabine, with or without capecitabine, fluorouracil with folinic acid, and the combination regimen FOLFIRINOX (fluorouracil, folinic acid, irinotecan, and oxaliplatin) (Petrelli *et al*., 2023). It has been suggested that combinatorial therapy against multiple targets may be more effective against cancer (Xelwa *et al*., 2021).

The tumour microenvironment (TME) of PDAC is characterised by a dense extracellular matrix (ECM) and several cell types (Kalluri, 2016; Jena *et al*., 2020). The crosstalk between numerous cell types, including cancer-associated fibroblasts (CAFs), myofibroblasts, tumour-associated macrophages (TAMs) and stromal cells plays a key role in tumour progression and in inducing chemoresistance (Shree *et al*., 2011; Feig *et al*., 2012; Shibuya *et al*., 2014; Ireland *et al*., 2016; Ireland and Mielgo, 2018; Schnittert, Bansal and Prakash, 2019; Jena *et al*., 2020). Unpacking the heterogeneity of PDAC TME is essential to understanding the dynamics of this disease and how cellular interactions, mediated by autocrine and paracrine factors, can induce signalling pathways that confer aggressive properties onto pancreatic cancer cells and foster chemoresistance.

SPP1 is a glycoprotein that is secreted by a variety of cell types, including osteoblasts, macrophages, T cells, fibroblasts, and dendritic cells (Del Prete *et al*., 2019), and has been linked to several physiological processes, such as bone resorption, wound healing, immune function, angiogenesis, cell survival, and cancer biology (Luo *et al*., 2015). One study suggests that *SPP1* may hinder apoptosis and facilitate the advancement of lung cancer cells. Increased *SPP1* expression in tumour tissue and plasma is a predictive marker associated with poor outcomes in several cancers (Cao *et al*., 2012; Chen *et al*., 2022). *SPP1* activation in stromal cells promotes cell survival and enhances the invasion of prostate cancer cells (Pang *et al*., 2019). Elevated expression of *SPP1* was found to be associated with poor clinical outcomes in patients with breast and ovarian cancer (Tu, Chen and Fan, 2019), suggesting that *SPP1* may be implicated in tumour growth and progression. However, the function of *SPP1* in PDAC with respect to tumour progression and chemoresistance remains to be fully elucidated.

This study aimed to demonstrate the gene expression patterns of signalling pathways in tumours of PDAC patients and identify potential targets in the patient group. *SPP1* was identified to be highly dysregulated. Subsequently, *SPP1* knockdown alone and in combination with gemcitabine was conducted to investigate the effects on pancreatic cancer cells.

## Materials and Methods

### Patient recruitment, study population and ethics

Ethics approval was obtained from the Human Research Ethics Committee (HREC) of the University of the Witwatersrand (Ethics number: M190735). After informed consent, tissue samples were collected from fifteen patients (15 tumour and 15 corresponding normal tissues) who underwent Whipple procedures at the Hepatopancreaticobiliary (HPB) units at Chris Hani Baragwanath Academic Hospital (CHBAH). The inclusion criteria included patients between the ages of 18 and 90 years and patients of African ancestry who had been histologically diagnosed with only PDAC. Patients undergoing chemotherapy were excluded. The recruited patients were aged between 53 and 95 years and included 7 females and 8 males (Nweke *et al*., 2020). These specimens were promptly submerged in RNA later™ stabilisation solution (Invitrogen™, Massachusetts, United States) for preservation and stored in a freezer until required.

### RNA extraction and cDNA synthesis

Tissues weighing between 15 to 25mg were dissected into small fragments using sterile surgical blades. These fragments were then placed into 15ml corning Falcon tubes (Thermo Fischer Scientific, Waltham, MA, USA) containing 700 μl of TRIzol reagent (Sigma Aldrich, Missouri, USA), and subsequently homogenized using a Tissue Ruptor (Qiagen, Hilden, Germany) then incubated at room temperature for 5 minutes. About 150µl of chloroform (Merck, Darmstadt, Germany) was added to each sample, mixed vigorously, and centrifuged at 12 000 x g at 4°C for 15 minutes, forming three layers. The transparent aqueous layer was transferred into a new labelled 1.5 ml centrifuge tube, with 350 μl of isopropanol (Sigma Aldrich St. Louis, Missouri, United States) added to the sample. The samples were resuspended, allowed to stand for 8 minutes at room temperature and centrifuged at 12 000 x g at 4°C for 8 minutes. The resulting supernatant was discarded, and the pellets were washed with 700 μl of 75% ice-cold ethanol (Sigma Aldrich St. Louis, Missouri, United States), and centrifuged at 7500 x g at 4°C for five minutes. Supernatants were discarded, and pellets were air-dried for 5 minutes. Pellets were resuspended in 30µl of nuclease-free water (Sigma Aldrich St. Louis, Missouri, United States), inserted into a heating block at 55-60°C for 5 minutes, and stored on ice. RNA purity was confirmed using the Nanodrop® 1000 (Thermo Fisher, Waltham, Massachusetts, United States) ensuring that the A260/A280 and A260/230 of the samples were above 2.0.

For cDNA synthesis, the RT^2^ cDNA synthesis kit (Qiagen, Hilden, Germany) was used. Tumour and normal tissue samples were pooled and adjusted to a concentration of 500ng/µL. A genomic DNA elimination mix was prepared, thoroughly mixed, and incubated at 42°C for 5 minutes using the SimpliAmp™ thermocycler (Thermo Fischer Scientific, Waltham, MA, USA) followed by a 1-minute cooling step on the ice.

### Targeted PCR arrays (Human Growth Factor and Signal Transduction)

For this study, the Human Growth Factor (PAHS – 041Z) and Signal Transduction arrays (PAHS – 014Z) from QIAGEN were used. According to the manufacturer’s instructions, 102 µL of cDNA was added to the PCR mixture and loaded onto the PCR arrays. Quant Studio 1 was used to amplify samples (Applied Biosystems, California, USA). The PCR reaction was run according to the manufacturer’s instructions. The QIAGEN GeneGlobe online software tool (https://geneglobe.qiagen.com/za/analyze/).

### Selection of target SPP1 and verification using real-time polymerase chain reaction

Following the screening of genes using the RT^2^ Profiler PCR Arrays, *SPP1* was selected. This selection was made because it was one of the most upregulated genes. *MRPL19* was used as the reference gene for the quantification of *SPP1* gene expression in tissue samples. An extensive literature search conducted also showed little information on the role of *SPP1* in PDAC especially in the context of chemoresistance, although it has been identified to be involved in key processes linked to carcinogenesis (Chen *et al*., 2022).

Real-time PCR was conducted on the same patient samples according to the MIQE guidelines (Bustin *et al*., 2009). The PCR reaction mix was prepared using the TaqMan® Fast Advanced Master Mix (Thermo Fischer, Massachusetts, USA). Real-time PCR was performed using primers designed in-house. The samples were run on a QuantStudio 1 Real-Time PCR system (Thermo Fischer Scientific, Waltham, MA, USA).

### Cell culturing of MIA PaCa-2 cells and drug treatment

Tumourigenic Human Pancreatic Carcinoma Cells (MIA PaCa-2) obtained from the Japanese Collection of Research Bioresources (JCRB) (Cat no JCRB0070), passage 2 (P2) was used in this study. The cell line underwent STR testing for confirmation.

The MIA PaCa-2 cell line was grown in HyClone^R^ DMEM/High Glucose growth medium (Hyclone, GE Life Sciences, Massachusetts, USA), enriched with 10% fetal bovine serum (FBS) (Biowest, Nuaillé, France) and 1% antibiotic (penicillin/streptomycin) (Thermo Fisher Scientific, Massachusetts, USA). The cells were kept at 37°C in a 5% CO_2_ incubator and were sub-cultured near confluency (±80%) every second day of the week. This was done by replacing the old growth medium with a fresh growth medium after rinsing twice with 1X PBS. Any excess sub-cultures were preserved at least −80°C in DMEM growth medium, supplemented with 70% FBS and 20% DMSO (dimethyl sulfoxide) (Thermo Fisher Scientific, Massachusetts, USA).

For treatment with gemcitabine, the MIA PaCa-2 cells were allowed to adhere for 24 hours before being exposed to control and test compounds diluted in DMEM. Briefly, the cells were exposed to different concentrations of gemcitabine (Sigma-Aldrich, St. Louis, MO, USA) (85, 42.5, 21.25, 10.63, 5.31, 2.66, 1.33, and 0.66 µM). Approximately 0.33 % DMSO was used as the vehicle control, and 0.4 µM doxorubicin (DOX) was used as the positive control drug, these concentrations were previously determined and employed in our laboratory. Each treatment was done in triplicate.

### siRNA knockdown of SPP1

Transfection of MIA PaCa-2 cells was conducted using *SPP1*-specific siRNA to investigate the potential of *SPP1* downregulation as a therapeutic strategy for the targeting of PDAC. Consequently, all subsequent assays were conducted post-transfection of MIA PaCa-2 cells with *SPP1*-specific siRNA (*Silencer*™ Select si-*SPP1*, Thermo Fisher Scientific, Massachusetts, USA), using the non-targeting control (NTC) siRNA (Silencer™ Select Negative Control #1 siRNA, Thermo Fisher Scientific, Massachusetts, USA) as negative control and *ACTB*-specific siRNA positive control (*Silencer*™ Select *ACTB* Positive Control siRNA, Thermo Fisher Scientific, Massachusetts, USA). Transfections were executed using the Lipofectamine 3000 Transfection Reagent (Thermo Fisher Scientific, Massachusetts, USA), adhering to the manufacturer’s protocol. Briefly, MIA PaCa-2 cells were seeded into 12-well plates and incubated overnight until they reached 60-70% confluency. The transfection reagent was mixed with pre-warmed DMEM only in a volume equivalent to that of the *SPP1* siRNA: (10nM siRNA: 10nM Lipofectamine 3000 transfection reagent), and NTC siRNA was then mixed separately with pre-warmed DMEM serum-free culture media under the same conditions.

The transfection reagent mixtures were combined with the corresponding siRNA mixtures, (10nM siRNA: 10nM Lipofectamine 3000 transfection reagent) and (10nM NTC siRNA: 10nM Lipofectamine 3000 transfection reagent) and they were subjected to a 15-minute incubation at room temperature to facilitate the formation of complexes. The complete solution of siRNA and transfection reagent were subsequently introduced into the respective wells, achieving a final volume of 1000 µl in the growth medium. The cells were then incubated for 24-48 hours. The transfected cells were imaged using the Olympus iX51 phase contrast microscope (Wirsam Scientific & Precision Equipment Ltd, Johannesburg, South Africa).

### Cell morphology, proliferation, migration assays

To evaluate the nuclear morphology of MIA PaCa-2 cells treated with gemcitabine, DAPI staining was utilized. The cells were seeded at a density of 1 x 10^5^ cells/well in a 12-well plate (Thermo Fischer Scientific, Waltham, MA, USA) in a complete medium at 37°C for 24 hours. Following 48 hours of treatment with 10 µM gemcitabine, the cells were washed twice with PBS and then fixed with 450 µl of 4% formaldehyde for 5 minutes. Next, the cells were permeabilized with 450 µl of 0.01% Triton for 5 minutes and then stained with five µg/ml of DAPI for 15 minutes in the dark. The cells were rewashed with PBS and examined under the Olympus iX51 fluorescent, inverted microscope (Wirsam Scientific & Precision Equipment Ltd, Johannesburg, South Africa). The images were visually scrutinized for morphological nuclear alterations suggestive of apoptosis.

The MTT (Sigma Aldrich, St. Louis, Missouri, United States), assay was employed to evaluate the viability of MIA PaCa-2 cells. This assay was performed on MIA PaCa-2 cells that were treated with varying concentrations of gemcitabine (0.66-85 µM) at 24 and 48 hours. The assay aimed to determine whether gemcitabine affected cell viability and to identify the optimal concentration that was toxic to the MIA PaCa-2 cells. The IC50 value, representing the concentration at which compounds inhibit cell viability by 50%, signifies the half-maximal inhibitory concentration. The assay relied on the colourimetric response generated by the presence of NAD(P)H-dependent oxidoreductases within the cells, which served as an indicator of metabolic activity and cell viability. Mitochondrial Succinate Dehydrogenase, a component of viable cells, reduced the tetrazolium dye, MTT, to its insoluble form, leading to the formation of purple formazan crystals. The quantity of crystals formed directly indicated cellular viability and provided a means of quantifying cell viability relative to an untreated sample. The MIA PaCa-2 cells were seeded into 96-well plates and incubated overnight in a humidified incubator (95% humidity, 37 ◦C and 5% CO2). MIA PaCa-2 cells were then treated using DMSO as a diluent control. Following incubation with the specific treatment concentration, 100 μl of 1 mg/ml MTT was added to each well and incubated at 37°C for 3 hours. The formazan crystals were dissolved in 200 μl of DMSO/well and the absorbance was measured at 450 nm using a spectrophotometer (MULTISKAN Sky, ThermoFisher Scientific) with a reference wavelength of 690 nm. The percentage cell viability was determined by using the treated values as a percentage relative to the corresponding untreated values for each biological triplicate.

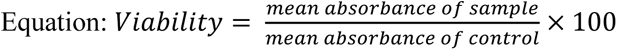

The purpose of the scratch assay was to evaluate the influence of *SPP1* knockdown, gemcitabine and combinational treatment on the migration capacity of MIA PaCa-2 cells. The cells were transfected as described, at 1 x 10^5^, in 96 well plates, and allowed to reach a confluency of 90-95%, ensuring the formation of a confluent monolayer. A vertical scratch was created across the cell layer using a P10 pipette tip, and markings were placed along the scratch on the underside of the plate for uniformity in the subsequent imaging procedure. The individual wells were imaged using an Olympus iX51 inverted microscope at 0 hour, and the cells were further imaged at 24, 48, and 72 hours post-scratch. The ImageJ software (National Institutes of Health and LOCI, University of Wisconsin) was used for image analysis of the scratch, which was expressed as a percentage of wound closure.

### Apoptosis assay

The apoptosis assay was carried out to investigate the effects of *SPP1* knockdown, gemcitabine treatment, and the combination of both on apoptosis in MIA PaCa-2 cells. The assay made use of the Annexin V-FITC staining kit (BD Biosciences, New Jersey) to detect early apoptosis, when phosphatidylserine is translocated to the outer leaflet of the plasma membrane is tranlocated. MIA PaCa-2 cells were seeded in 12-well plates at a density of 2 x 10^5^ cells/well for 24 hours and were then transfected with siRNA targeting *SPP1*. The cells were further treated with 10 µM gemcitabine for 48 hours, after which they were washed with PBS and trypsinized. Next, 200 µl of complete media was added to inactivate the trypsin. The cells were then centrifuged at 1500 x g for 5 minutes at 4°C, after which they were resuspended in 2 ml of complete medium and incubated for 30 minutes to allow recovery from trypsinization. The cells were washed with 2 ml of PBS, centrifuged at 1200 x g for 5 minutes, and resuspended in 100 µl of 1X binding buffer (from the BD Annexin V/FITC Apoptosis kit). The suspensions were transferred into flow tubes and stained with 2 µl of Annexin V and 2 µl of propidium iodide (PI). Brief vortexing and incubation of the cells for 15 minutes at room temperature in the dark followed. Subsequently, 250 µl of 1X binding buffer was added to each tube, and the cells were analyzed using a flow cytometer. Data acquisition was undertaken at low flow rates, which were less than 300 events per second, using the BD LSRFortessa™ Analyser and FACSDiva software (BD Biosciences, Franklin Lakes, NJ, USA). This was done to achieve enhanced resolution and precise DNA quantification. A minimum of 30,000 events were recorded per sample, and the fluorescent signal from the samples was detected using a 610/20 band-pass filter. The generated flow cytometry standard (FCS) files in FACSDiva were then imported and analysed using FlowJov10.8.1 (FlowJo, LLC, Ashland, KY, USA).

### SWATH-MS proteomics analysis

After pelleting the cells, they were resuspended in a 50 mM Tris-HCl pH 8.0 solution and 2% SDS. The Pierce Bicinchoninic assay was used to measure protein concentration as per the manufacturer’s instructions (Thermo Fisher Scientific). To prepare the protein samples for analysis, each sample containing 10 μg of protein underwent a single-step reduction and alkylation using 5 mM tris (2-carboxyethyl) phosphine and 10 mM 2-chloroacetamide, at room temperature for 20 minutes. Subsequently, the samples were subjected to a purification process, removing detergents and salts, utilizing MagReSyn™ HILIC beads (ReSyn Biosciences) as previously described (Nweke *et al*., 2020; Baichan *et al*., 2023). On-bead protein digestion was carried out with a 1:20 ratio of protease to protein, employing sequencing-grade trypsin. The resulting peptides were dried and preserved at −80°C until they were ready for LC-MS analysis.

Samples, with about 1.5μg of peptides each, underwent analysis via a Dionex Ultimate 3000 RSLC system linked to a Sciex 5600 TripleTOF mass spectrometer. An Acclaim PepMap C18 trap column (75 μm × 2 cm; 2 min at 5 μl.min 1 using 2% ACN/0.2% FA) was used to inline de-salt injected peptides. Trapped peptides were gradient eluted and separated on a Waters Acquity CSH C18 NanoEase column (75 μm × 25 cm, 1.7 µm particle size) at a flow rate of 0.3 µl.min 1 with a gradient of 6-40% B over 90 min (A: 0.1% FA; B: 80% ACN/0.1% FA). For Sequential window acquisition of all theoretical mass spectra (SWATH), precursor scans were acquired from 400-1100 m/z with 50 milliseconds accumulation time, and fragment ions were acquired from 200-1800 m/z for 48 variable-width precursor windows with 0.5 Da overlap between windows and 20 milliseconds accumulation time per window.

SWATH data was processed using Spectronaut v17 software (Biognosys). For data processing, the default direct DIA identification and quantification settings were used. As a fixed modification, carbamidomethylation was added, and as variable modifications, N-terminal acetylation and methionine oxidation were added. Swiss-Prot human sequences (downloaded on 03 March 2023 from www.uniprot.org) and common contaminating proteins were used as the search database. A q-value ≤ 0.01 cut-off was applied at the precursor and protein levels. Quantification was performed at the MS1 and MS2 levels. Label-free cross-run normalization was employed using a global normalization strategy. Candidate dysregulated proteins were filtered at a q-value ≤ 0.01, absolute Log2 fold change (FC) ≥ 1, and a minimum of two unique peptides were identified.

### Pathway and statistical analyses

To evaluate the dysregulated protein sets, functional enrichment analysis and network analysis using Cytoscape v3.8.2 were performed (Shannon *et al*., 2003). The stringApp v2.0.1 (Doncheva *et al*., 2019) within Cytoscape to examine the molecular functions, cellular compartments, and pathways (Reactome v8.0.5 (Fabregat *et al*., 2017) and KEGG (Kanehisa *et al*., 2016) databases) associated with the dysregulated proteins. Query parameters included Homo sapiens as the reference species, a confidence threshold of 0.4, and no inclusion of additional interactors. The Shapiro–Wilk test was conducted to test for data normality. Subsequently, an unpaired Student t-test was employed for comparison between the two groups. The non-parametric Kruskal–Wallis Anova by Ranks was performed, followed by multiple comparisons using the Benjamini – Hochberg test. A p-value of less than 0.05 was considered statistically significant.

## Results

### Differentially expressed genes and pathways in tumours

To demonstrate the differentially expressed genes in PDAC tissues, two RT^2^ PCR array panels were used. The results showed that from the growth factor array panel, 48 were upregulated and 10 were downregulated in tumours compared to adjacent normal tissues (Supplementary Table 1). For the signal transduction array panel, a total of 56 genes were found to be upregulated and 10 genes were downregulated (Supplementary Table 1). After a screening of the dysregulated genes from both panels, *SPP1* was selected as a gene of interest. Consequently, qPCR was applied to verify the expression of *SPP1* in PDAC tumour tissues and corresponding adjacent normal tissues. The expression of *SPP1* was significantly elevated in tumour tissues compared to normal tissues (*p* < 0.0001).

**Figure 1:**
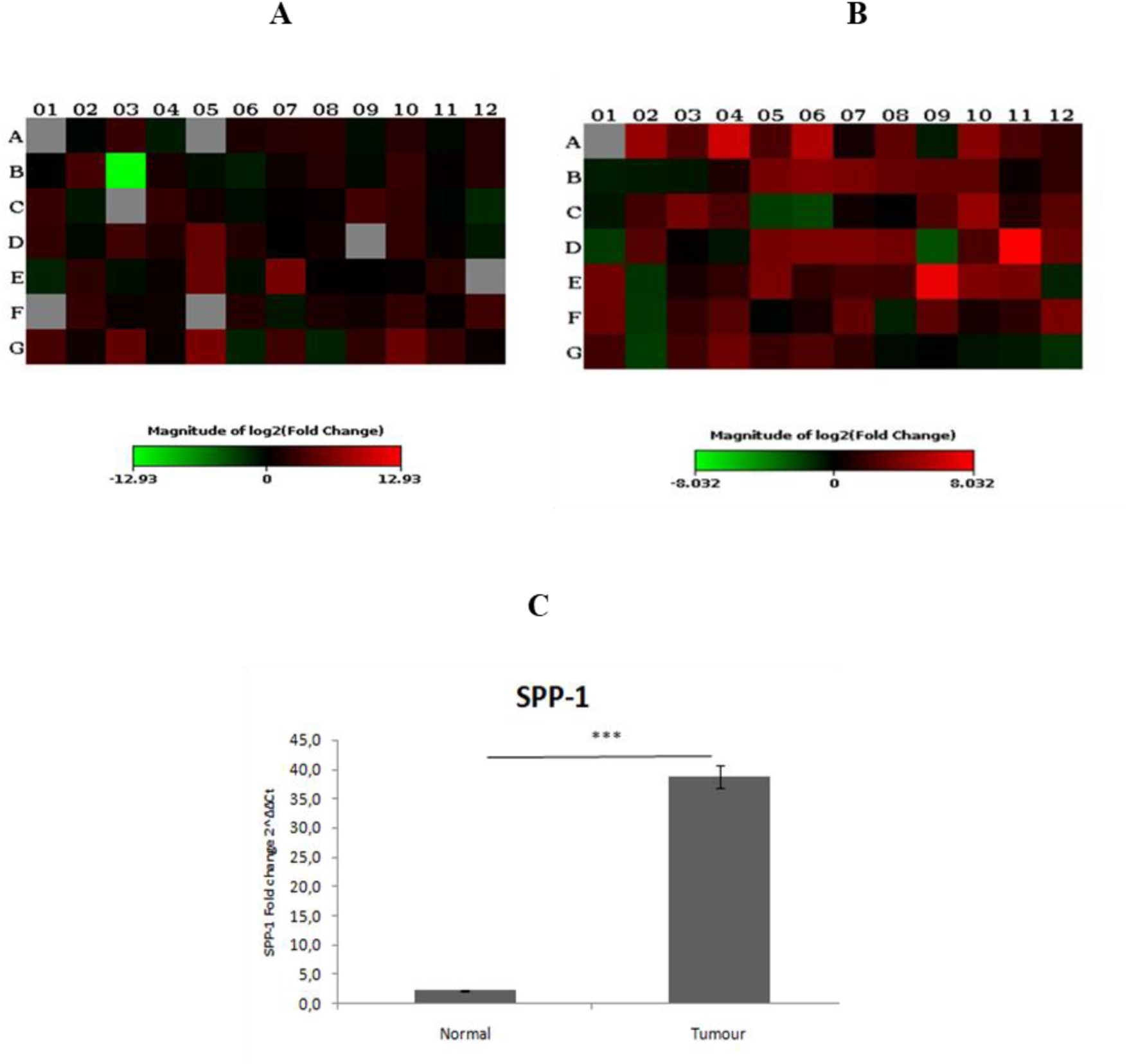
*SPP1* overexpression in tissues of PDAC patients using real-time PCR. Heat map representation of the fold changes in expression for every gene in the array. (A) 48 were found to be upregulated, and 10 were downregulated in tumours compared to adjacent normal tissues in the Growth factor array. (B) 55 genes were found to be upregulated, and 10 were downregulated in tumours compared to adjacent normal tissues in the Signal Transduction array. **(C)** Quantification of *SPP1* gene expression in tissue samples was performed using real-time PCR. p values less than 0.05 were considered statistically significant (*** denotes *p* < 0.001).

To shed light on the network and pathway interactions present in the dataset the Reactome pathway browser was used to interrogate the dysregulated genes and identify enriched pathways, respectively. The genes upregulated and downregulated from both panels were combined to identify key upregulated and downregulated pathways. The top upregulated pathways included those associated with interleukin signalling (*IL4* and *IL13*), cytokine signalling, receptor kinase signalling, and PI3/Akt signalling. The downregulated pathways included IL-10 signalling, immune system and several transcription regulation pathways. Furthermore, network analysis using String demonstrated an intricate network for the upregulated and downregulated genes suggesting the genes interact together and are involved in similar biological processes and pathways.

### SPP1 knockdown decreases cell migration potential in MIA PaCa-2 cells

Cell morphology was assessed using brightfield, phase contrast microscopy (Olympus iX51 inverted microscope), following a 48-hour treatment with 10 µM of gemcitabine, as well as after *SPP1* knockdown. *SPP1* knockdown was confirmed by real-time PCR (Supplementary Fig. 1). A combination of *SPP1* knockdown and gemcitabine treatment showed a drastic change in the morphology of the cells following 96 hours of treatment (Figure 2a). The observation of cell shrinkage and detachment, which are characteristic of apoptosis (Ziegler and Groscurth, 2004), although not conclusively. To investigate the influence of *SPP1* knockdown and combination treatment with gemcitabine on cell migration of MIA PaCa-2 cells, the wound healing assay or scratch assay was used. A ‘wound’ was created in cells with a confluence of 90-95%, which were then treated with either *SPP1* knockdown alone, gemcitabine treatment alone, or a combination of both *SPP1* knockdown and gemcitabine treatment. The closure of the wound was then observed at 24 hours, 48 hours, and 72 hours post-treatment (Figure 2b). At 24, 48, and 72 hours, the untreated and diluent control exhibited an increase in wound closure. Of note, gemcitabine treatment resulted in an increase in closure rate at 72 hours compared to *SPP1* knockdown alone and combination of *SPP1* knockdown and gemcitabine treatment. The migration of MIA PaCa-2 cells was markedly inhibited upon *SPP1* knockdown, at all time points. Furthermore, the combination of *SPP1* knockdown and gemcitabine treatment resulted in inhibition of MIA PaCa-2 cell migration, compared to untreated, diluent control, *SPP1* knockdown alone and gemcitabine alone (Figure 2c).

**Figure 2:**
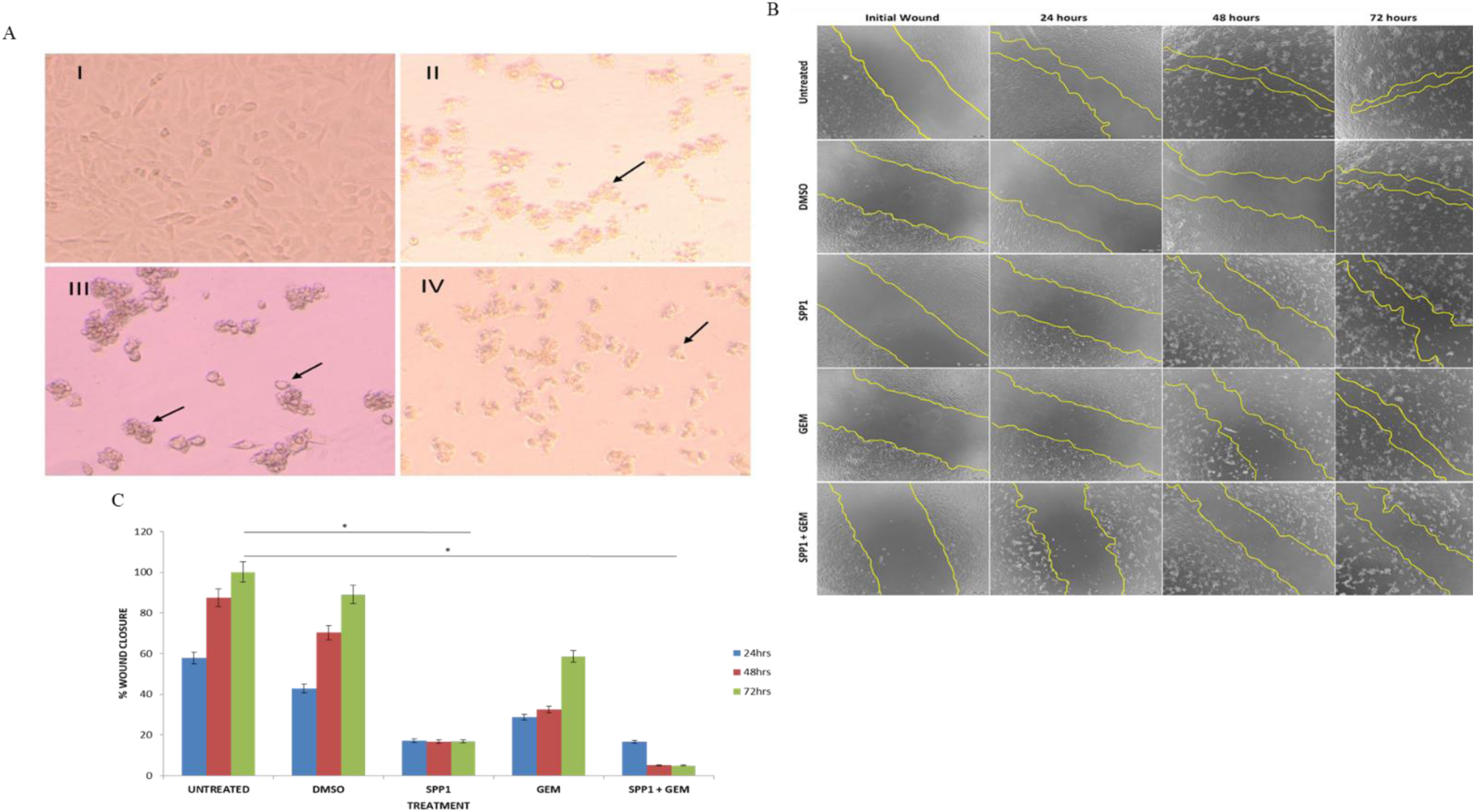
Combination of *SPP1* knockdown and gemcitabine treatment decreases cell migration of MIA PaCa-2 cells and induces apoptotic morphology. (A) A phase contrast microscope was used to assess cellular morphological changes following *SPP1* knockdown, gemcitabine treatment, and a combination of *SPP1* knockdown and gemcitabine treatment. Representative phase-contrast images (I-IV). MIA PaCa-2 cells that were (I) untreated, (II) 48-hour treatment with 10 µM of gemcitabine (III) *SPP1* knockdown for 48 hours (IV) Combination treatment of *SPP1* knockdown and gemcitabine treatment at 96 hours. The combination of *SPP1* knockdown and gemcitabine treatment significantly decreases cell migration of MIA PaCa-2 cells following treatment for 48 hours, 72 hours. (B) The migration abilities of MIA PaCa-2 cells were determined by scratch assay, the regions highlighted in yellow represent the wound area. (C) Graph representation of measured wound closure area. * denotes *p* < 0.05.

### Combination of SPP1 knockdown and gemcitabine treatment induces apoptosis

To investigate the potential synergistic induction of apoptosis, MIA PaCa-2 cells underwent *SPP1* knockdown combined with gemcitabine treatment for 96 hours. Thereafter flow cytometry analysis was conducted to determine whether apoptosis was a cause of cell death in MIA PaCa-2 cells.

Cells were assessed for apoptosis (Annexin-V+/PI+) (Figures 3a and 3b). As expected gemcitabine induces early apoptosis in MIA PaCa-2 cells (24.5%), being the first line of treatment for PDAC, only marginally less than the positive control (DOX). Interestingly, *SPP1* alone did not show any significant apoptosis induction in MIA PaCa-2 cells following knockdown, with 5.7% of cells being in early apoptosis compared to 2.5% of untreated cells. Importantly, the combined treatment demonstrated a heightened apoptotic effect compared to the application of each treatment individually. *SPP1* knockdown significantly increased gemcitabine’s apoptosis-promoting impact, demonstrating a synergistic effect (37%).

**Figure 3:**
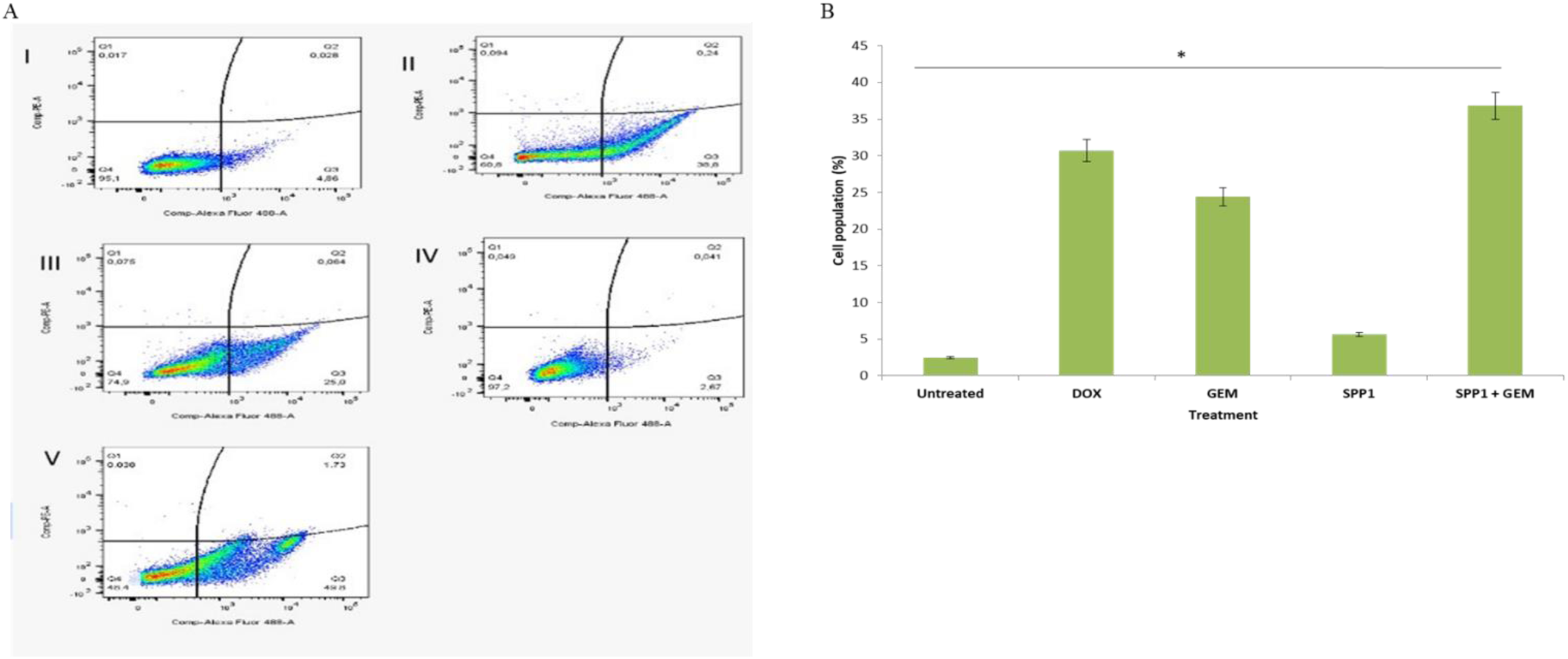
Effect of *SPP1* knockdown only, gemcitabine treatment only, and the combination on apoptosis using PI and Annexin V. (**A**) Cell apoptosis analysis demonstrated with the quadrants (Q1: cells undergoing secondary necrosis (PI+); Q2: cells undergoing late stage apoptosis (Annexin V+ PI+); Q3: cells undergoing early apoptosis (Annexin V+); Q4: live cells. All subsequent graphs show cells stained with Annexin-V and PI. (I) Untreated cells. (II) DOX (positive control) (III) gemcitabine treatment only. (IV) *SPP1* knockdown only. (V) *SPP1* knockdown and gemcitabine treatment. (**B**) Graph representation of cells in early apoptosis (Annexin V positive and PI negative double-staining). *SPP1* knockdown significantly enhanced the apoptosis-promoting effect of gemcitabine. * denotes a p-value of less than 0.05.

### Proteomic profiling of treated cells reveals dysregulated proteins

A comprehensive proteomic analysis was conducted to further gain an understanding of *SPP1*’s role in pancreatic cancer and to provide deeper insights into the pathways affected by *SPP1* downregulation. The profile of dysregulated proteins for the various treatments (gemcitabine treatment, *SPP1* knockdown, and combination treatment) compared to untreated MIA PaCa-2 cells are described (Table 2.1, 2.2 and 2.3). In the gemcitabine-treated group, there were a total of 893 dysregulated proteins, of these 439 proteins were upregulated and 454 proteins were downregulated. In the *SPP1* knockdown-treated group, there were a total of 40 dysregulated proteins, 27 were upregulated and 13 were downregulated. The combination treatment group indicated that there were 188 proteins dysregulated compared to the untreated group. Of these, 110 proteins were upregulated and 78 proteins were downregulated. Thereafter, unique and commonly dysregulated proteins across treatments were elucidated using Venny (v2.1.0). There were 741 proteins unique to gemcitabine treatment, 13 proteins unique to *SPP1* knockdown treatment, 34 proteins unique to combination treatment, and 13 proteins which were common across all 3 groups (Figure 4).

**Table 1:**
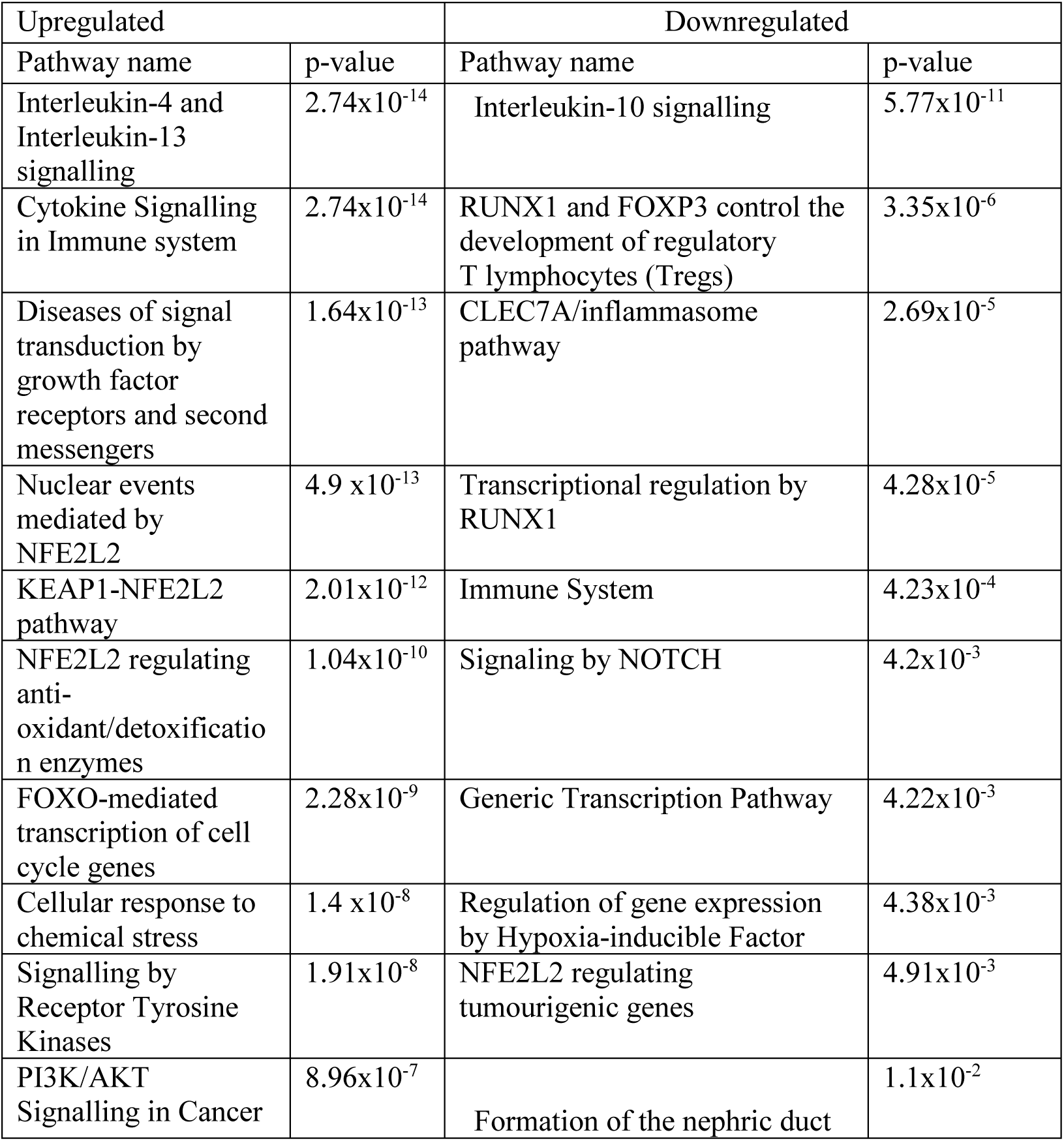
Pathways in which dysregulated genes are involved.

| Upregulated |  | Downregulated |  |
| --- | --- | --- | --- |
| Pathway name | p-value | Pathway name | p-value |
| Interleukin-4 and Interleukin-13 signalling | $2.74 \times 10^{-14}$ | Interleukin-10 signalling | $5.77 \times 10^{-11}$ |
| Cytokine Signalling in Immune system | $2.74 \times 10^{-14}$ | RUNX1 and FOXP3 control the development of regulatory T lymphocytes (Tregs) | $3.35 \times 10^{-6}$ |
| Diseases of signal transduction by growth factor receptors and second messengers | $1.64 \times 10^{-13}$ | CLEC7A/inflammasome pathway | $2.69 \times 10^{-5}$ |
| Nuclear events mediated by NFE2L2 | $4.9 \times 10^{-13}$ | Transcriptional regulation by RUNX1 | $4.28 \times 10^{-5}$ |
| KEAP1-NFE2L2 pathway | $2.01 \times 10^{-12}$ | Immune System | $4.23 \times 10^{-4}$ |
| NFE2L2 regulating anti-oxidant/detoxification enzymes | $1.04 \times 10^{-10}$ | Signaling by NOTCH | $4.2 \times 10^{-3}$ |
| FOXO-mediated transcription of cell cycle genes | $2.28 \times 10^{-9}$ | Generic Transcription Pathway | $4.22 \times 10^{-3}$ |
| Cellular response to chemical stress | $1.4 \times 10^{-8}$ | Regulation of gene expression by Hypoxia-inducible Factor | $4.38 \times 10^{-3}$ |
| Signalling by Receptor Tyrosine Kinases | $1.91 \times 10^{-8}$ | NFE2L2 regulating tumourigenic genes | $4.91 \times 10^{-3}$ |
| PI3K/AKT Signalling in Cancer | $8.96 \times 10^{-7}$ | Formation of the nephric duct | $1.1 \times 10^{-2}$ |

**Table 2.1:**
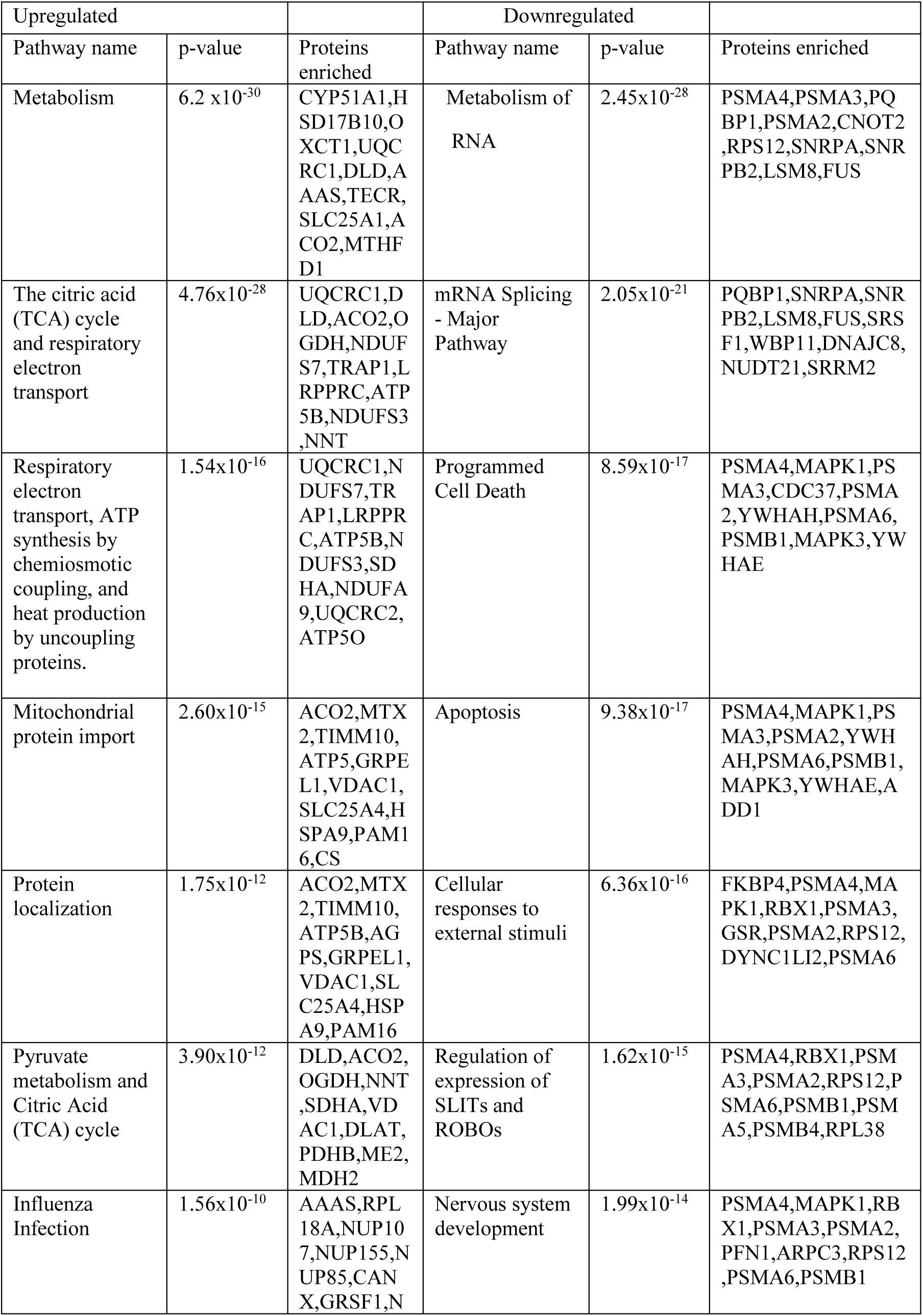
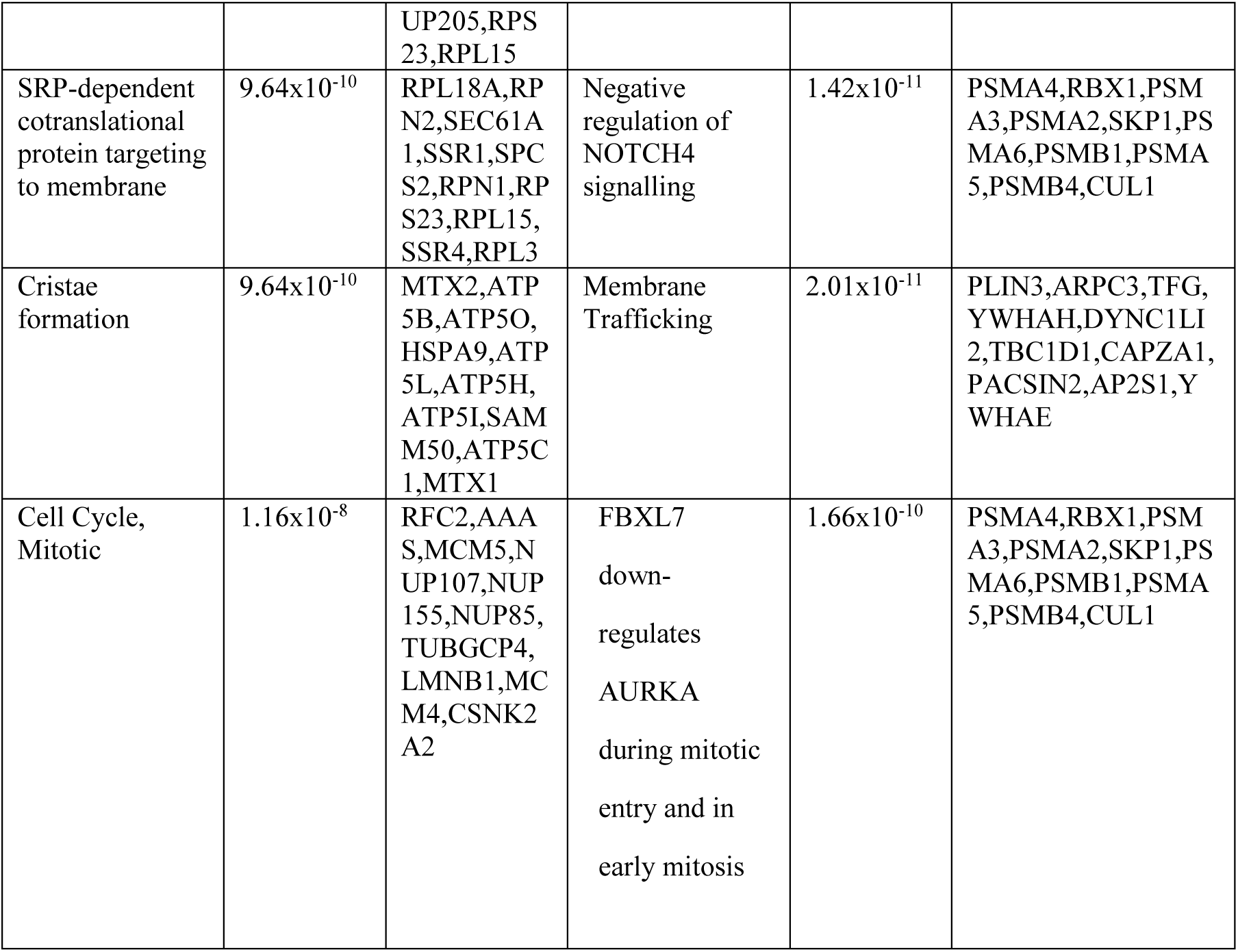
Dysregulated pathways following gemcitabine treatment.

**Table 2.2:**
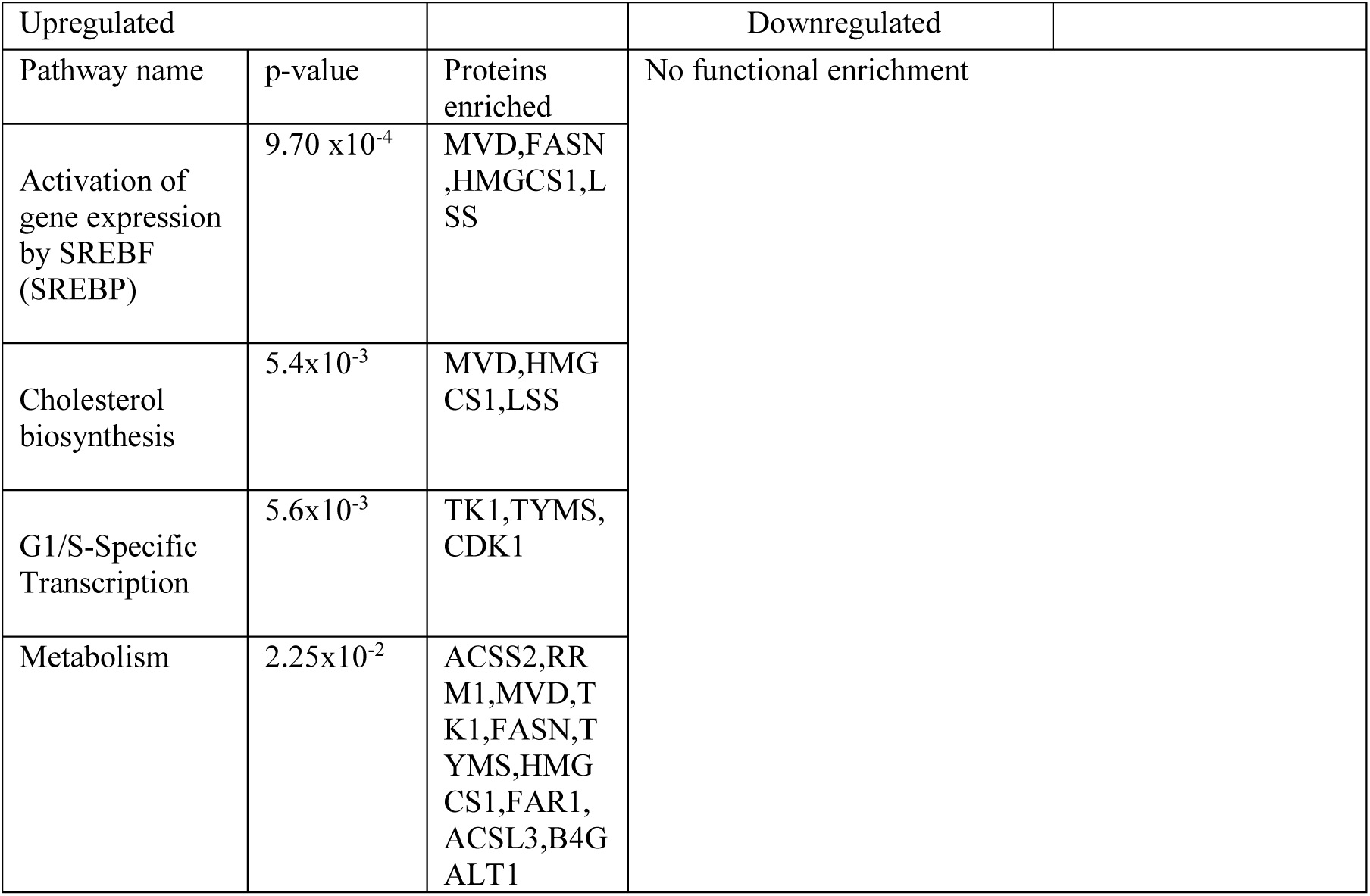
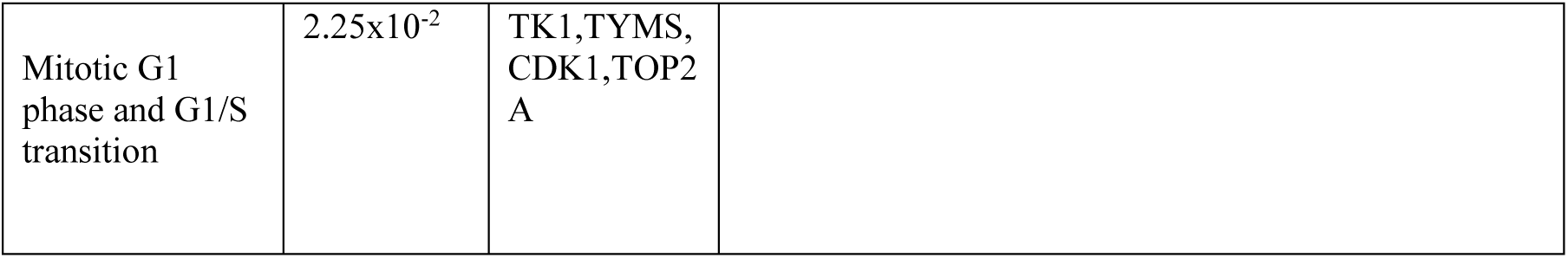
Dysregulated pathways following *SPP1* knockdown.

| Upregulated |  |  | Downregulated |
| --- | --- | --- | --- |
| Pathway name | p-value | Proteins enriched | No functional enrichment |
| Activation of gene expression by SREBF (SREBP) | 9.70 x10<sup>-4</sup> | MVD,FASN,HMGCS1,LSS |  |
| Cholesterol biosynthesis | 5.4x10<sup>-3</sup> | MVD,HMGCS1,LSS |  |
| G1/S-Specific Transcription | 5.6x10<sup>-3</sup> | TK1,TYMS,CDK1 |  |
| Metabolism | 2.25x10<sup>-2</sup> | ACSS2,RRM1,MVD,TK1,FASN,TYMS,HMGCS1,FAR1,ACSL3,B4GALT1 |  |

|  |  |  |
| --- | --- | --- |
| Mitotic G1 phase and G1/S transition | 2.25x10<sup>-2</sup> | TK1,TYMS, CDK1,TOP2 A |

**Table 2.3:**
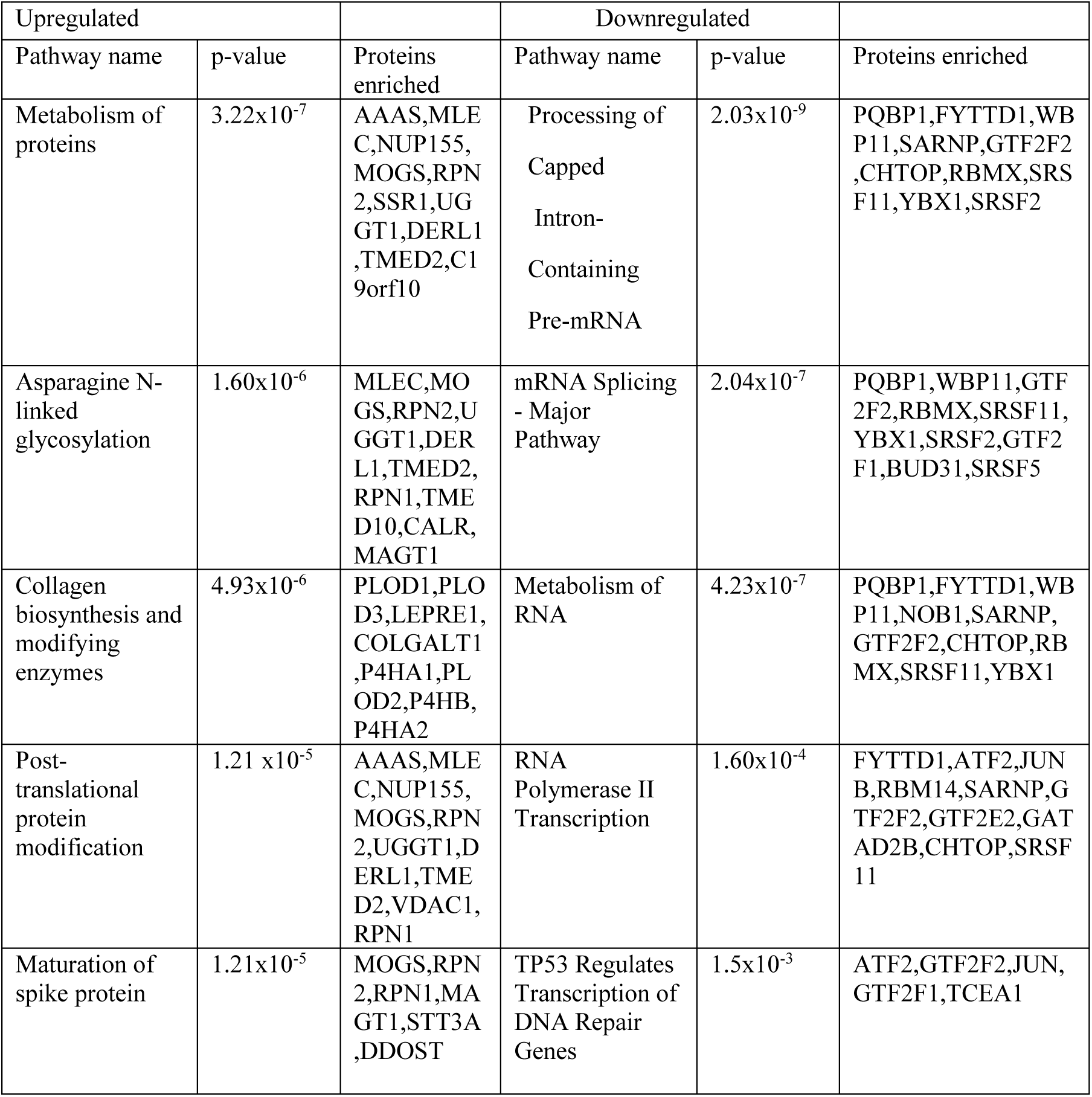
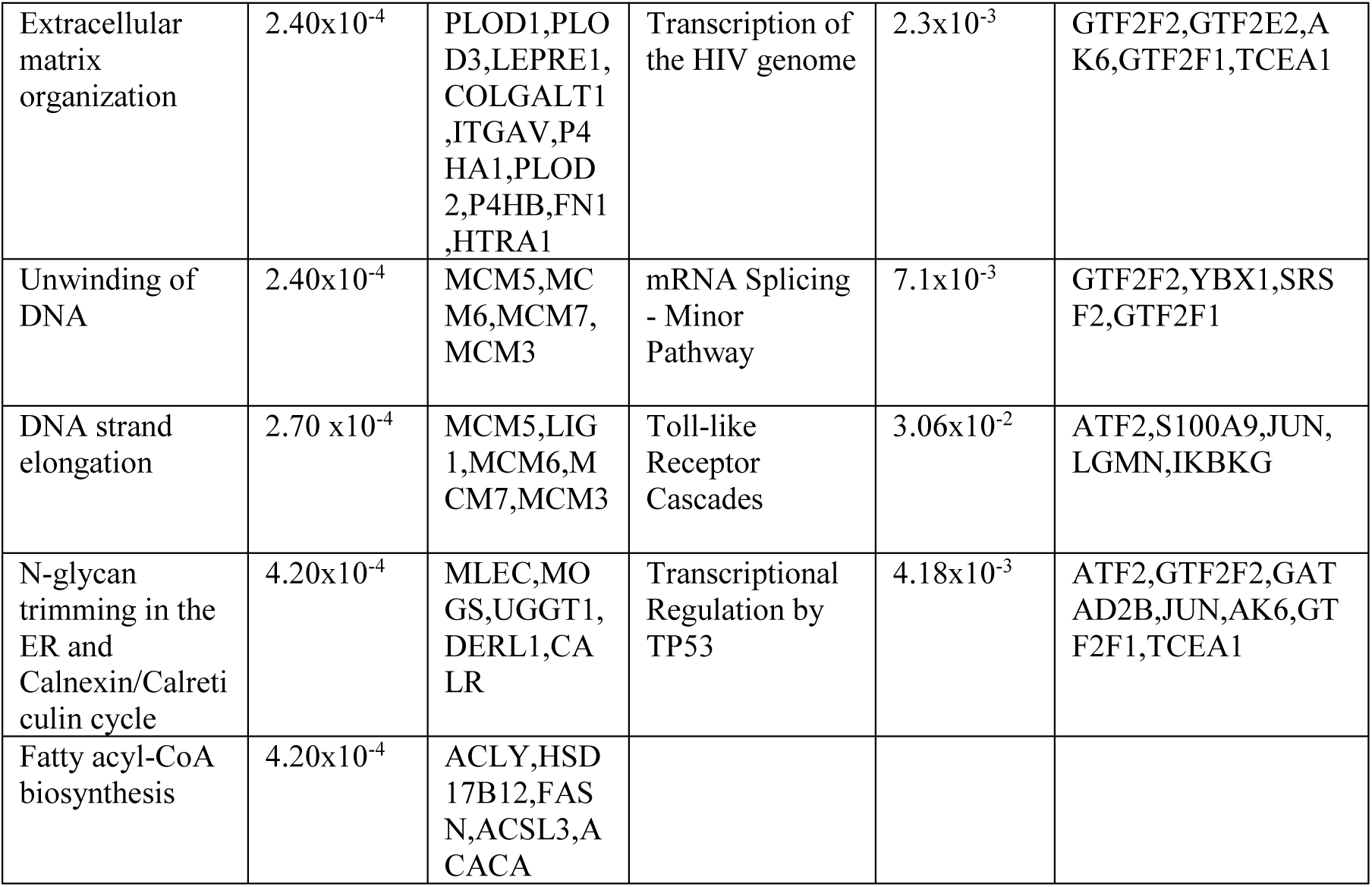
Dysregulated pathways following a combination of *SPP1* knockdown and gemcitabine treatment.

**Figure 4:**
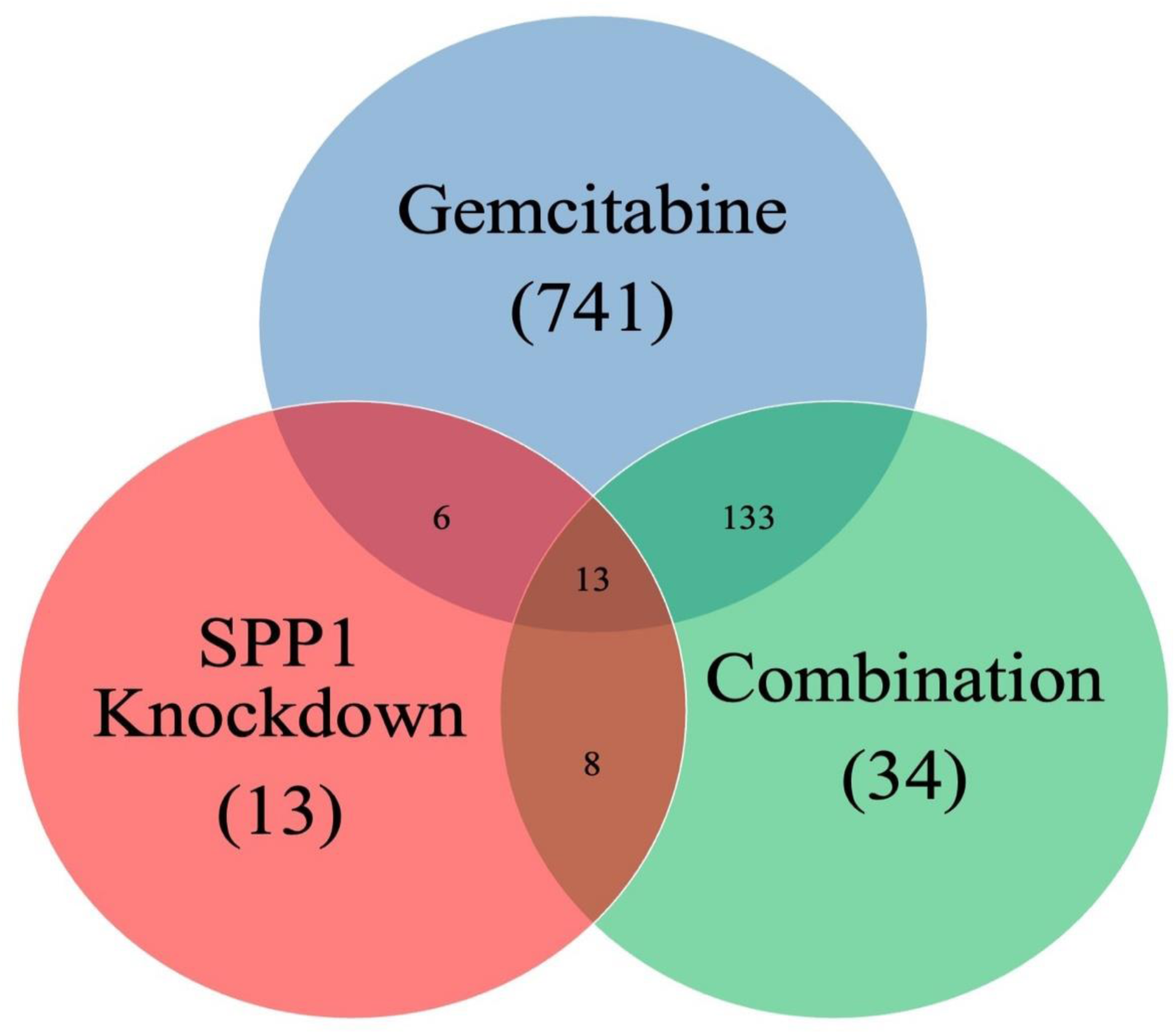
Venn diagram depicting uniquely and commonly dysregulated proteins. There were 741, 13, and 34 proteins uniquely dysregulated in the gemcitabine, *SPP1* knockdown, and combination treatment groups, respectively. Thirteen proteins were commonly dysregulated across all 3 treatment groups. Replicates were consistent across the treatment groups gemcitabine treatment, *SPP1* knockdown treatment, combination treatment, and untreated control

Principal component analysis (PCA analysis) was conducted to ascertain whether the differentially expressed proteins in each of the treatment groups were able to successfully distinguish between each other (Supplementary Fig. 2). PCA analysis shows that PC1 accounts for 63.92% variation and PC2 accounts for 10.05% variation. The gemcitabine treatment, *SPP1* knockdown, and combination treatment were distinguished along PC1. However, the *SPP1* knockdown and untreated sample groups overlap showing that the protein profiles of these two groups were similar; notwithstanding that the untreated samples cluster tightly together while the *SPP1* knockdown shows more variance across PC2.

### Aberrant pathways with SPP1 knockdown and combined treatment with gemcitabine

Pathway analysis on the differentially expressed proteins was performed to determine the specific processes the dysregulated proteins in gemcitabine treatment (Table 2.1), *SPP1* knockdown (Table 2.2), and combination treatment (Table 2.3) groups are involved in. Metabolism, including a member of the cytochrome p450 family CYP51A1, was a top pathway enriched by the dysregulated proteins in the gemcitabine and combination treatment groups. Proteins involved in the cell cycle were enriched in Gemcitabine treatment only, including MCM5 which is involved in the transition from G0/G1/S phase. Cells that underwent gemcitabine treatment alone and in combination with *SPP1* knockdown were enriched in proteins involved in apoptosis. In the *SPP1* knockdown group, transcription activation was one of the most enriched pathways by the differentially expressed proteins, with an upregulation of *CDK1,* a major player in cell cycle transition. Under a combination of both *SPP1* knockdown and gemcitabine treatment, not only were proteins involved in metabolism upregulated, but also proteins involved with collagen biosynthesis and extracellular matrix remodelling, including members of the *PLOD* family. Moreover, there was a downregulation of Toll-like receptor cascades and the upregulation of, for example, *STAT3* involved in particularly the adaptive immune response.

## Discussion

The architecture of the TME of PDAC contributes significantly to chemoresistance. Chemoresistance poses a significant challenge in managing PDAC (Yu, Zhang and Xie, 2021), and its complexity is compounded by the invasive and metastatic nature of the disease, which is further exacerbated by its high molecular heterogeneity propagated by various signalling pathways (Connor and Gallinger, 2022). The understanding of the interplay between these pathways is crucial in identifying novel treatment strategies.

In this study, we profiled the key genes in crucial signalling pathways and selected *SPP1* as a potential target. SPP1, a protein with multiple cellular functions, that is overexpressed in various types of cancer. It is involved in biological processes such as bone metabolism, immune regulation, and wound healing, as well as in the survival and progression of cancer cells (Zhang *et al*., 2020; Kiss *et al*., 2021). Its role and overexpression in PDAC tumours may hint towards its role in tumourigenesis suggesting it could be effectively targeted.

The study also revealed that decreasing the expression of *SPP1* via knockdown led to a decline in the migratory and invasive capabilities of highly aggressive PDAC cells. This is consistent with findings by Xu et al. suggesting that inhibiting the expression of *SPP1* through siRNA significantly hampers the ability of MIA PaCa-2 cells to migrate (Xu *et al*., 2017). Specifically, diminishing *SPP1* expression impedes migration, and invasion, while elevated *SPP1* expression is strongly associated with a dismal prognosis in various forms of cancer (Liu *et al*., 2020). This has been shown in breast cancer studies, with Zhang et al.’s findings which suggest that *SPP1* expression is associated with heightened cell migration and adhesion in breast cancer (Zhang *et al*., 2014); and Rizwan et al. who observed that the knockdown of *SPP1* in MDA-MB-231 breast cancer cells resulted in a significant decrease in migration during *in vitro* transwell migration assays (Rizwan *et al*., 2018). According to a study by Zeng et al. inhibiting *SPP1* in ovarian cancer was shown to decrease cell proliferation, migration, and invasion capacities through the β1/FAK/AKT signalling pathway (Zeng *et al*., 2018). This study corroborates the findings of others suggesting the role of *SPP1* in inhibiting cell migration and invasion.

Furthermore, we showed that the combination therapy of *SPP1* knockdown and gemcitabine treatment enhanced apoptosis in MIA PaCa-2 cells. This indicates a potential role of SPP1 in circumventing chemoresistance. This hypothesis is supported by previous studies in lung adenocarcinoma (Matsubara *et al*., 2022) and prostate cancer (Pang *et al*., 2021). Studies have shown that the expression of *SPP1* on cancer cells in lung cancer patients is associated with chemoresistance and poor prognosis (Yan *et al*., 2015; Ouyang *et al*., 2018; Matsubara *et al*., 2022). Additionally, studies by Wang et al. found that the SPP1-EGFR pathway promotes stem-like features and is also linked to the development of radiation resistance in *KRAS*-mutated lung cancer (Wang *et al*., 2017). Furthermore, lung adenocarcinoma tissues showed lower expression of E-cadherin and higher expression of *SPP1* in the EMT region while an in vitro study found that the EMT process initiated by *SPP1* was inhibited by PI3K/Akt and MAPK signalling suppressors (Shi *et al*., 2021). In castration-resistant prostate cancer overexpression of *SPP1* has a significant influence on the regulation of apoptosis, as studies by Pang et al., have shown that a decrease in *SPP1* levels can inhibit apoptosis through the PI3K/Akt signalling pathway (Pang *et al*., 2021). In certain cancers (prostate and colon), an increased correlation between activated PI3K/Akt and poor patient outcomes has been observed. Therefore, it is anticipated that reducing *SPP1* expression might decrease the activation of the PI3K/Akt signalling pathway, preventing the activation of survival pathways and promoting the initiation of apoptosis (Pang *et al*., 2021). Moreover, in prostate cancer cells, *SPP1* elevation of the expression of p-glycoprotein has been linked to the emergence of multidrug resistance (Hsieh *et al*., 2013). Similarly, the levels of *SPP1* expression in a cohort of breast cancer patients who underwent adjuvant chemotherapy were discovered to predict the effectiveness of neoadjuvant chemotherapy in certain cases (Insua-Rodríguez *et al*., 2018).

Despite the proportion of cells undergoing early apoptosis and the decrease in migratory capacity visualised at 48 hours, further proteomic analysis of the effects of chemotherapy and *SPP1* knockdown revealed induced compensatory mechanisms for survival. The expression profiling data following *SPP1* knockdown indicated an up-regulation of genes related to cell cycle (*TK1, TYMS, CDK1, TOP2A*), and *CDK1* is thought to contribute to the development of PDAC by facilitating cell cycle progression and resisting apoptosis. *CDK1* is a gene that plays a critical role in promoting the progression of PDAC cells through the cell cycle (Lim and Kaldis, 2013). The CDKs, including *CDK1*, are vital in cell-cycle regulation and transcriptional elongation, and their dysregulation has been implicated in the progression of various cancers, including PDAC (García-Reyes *et al*., 2018). The G1/S-specific transcription factors that were found to be upregulated despite *SPP1* knockdown, indicate different avenues by which MIA PaCa-2 cells can continue driving growth. *CDK1* regulates mitosis entry and progression, as well as collaborates with cyclins like D1, E, and A to manage the progression of the G1 phase and S phase transition (Novak *et al*., 2007). In response to stress or DNA damage, *TP53* inhibits *CDK1* signalling, which prevents cancerous cell growth through apoptosis. However, overexpression of *CDK1* can promote cell replication with DNA abnormalities, contributing to cancer cell proliferation. It has been established that the activity of CDK1 is essential for the development of tumours (García-Reyes *et al*., 2018). The expression of *CDK1* genes is significantly higher in tumour cells of PDAC patients, indicating advanced stages of PDAC and poorer survival outcomes (Dong *et al*., 2019).

Toll-like receptors (TLRs) appear to exhibit a dual function, as they can both promote and inhibit the progression of PDAC (Vaz and Andersson, 2014). TLR7 and TLR9 stimulation in either stromal or transformed epithelial cells has been reported to accelerate tumour progression (Ochi *et al*., 2012; Zambirinis *et al*., 2015). Studies have shown a significant increase in the expression of TLR2, TLR4, and TLR9 in PDAC (Grimmig *et al*., 2016; Nurmi *et al*., 2022; Topcu *et al*., 2022). Our study found that the TLR signalling pathway’s activity was downregulated, which may hinder PDAC cell growth by reducing inflammatory markers and promoting apoptosis, as shown by Ajay et al. (Ajay *et al*., 2024). The combinational treatment of *SPP1* knockdown and gemcitabine treatment could limit PDAC tumour growth by decreasing TLR 7 and TLR 9, which are involved in tumour development.

## Conclusion

The current study demonstrated key alterations in signalling pathways in a sample group of PDAC patients. *SPP1* was further verified as upregulated in PDAC tumours. siRNA knockdown of *SPP1* in conjunction with gemcitabine treatment in MIA PaCa-2 cells altered various hallmark traits associated with PDAC including a significant reduction in cell proliferation, an increase in apoptosis, and a decrease in migration capacity. This indicates that this combinatorial treatment may have synergistic effects against PDAC cells. Additionally, the findings from the proteomic analysis demonstrated the potential molecular mechanisms associated with the phenotypic observations following combinatorial treatment such as the downregulation of the Toll-like receptor cascades. Future studies need to elucidate some of these mechanisms, especially concerning other compensatory mechanisms identified during the combined treatments and the effect of this treatment in *in vivo* models.

## Authors’ contributions

E.E.N conceptualised the study. N.X, P.N, G.C, and E.E.N acquired funding for the project. N.X, P.N, and E.E.N collected data. N.X, P.N, J.D, J.OJ, M.S, T.N.A, G.C, and E.E.N performed data analysis and interpretation. N.X, P.N, T.N.A and E.E.N wrote the initial draft. All authors critically reviewed and approved the final manuscript.

## Ethics clearance

Ethical clearance for this study was obtained from the Human Research Ethics Committee (Medical) of the University of the Witwatersrand (M190735). Before sample collection, all participating individuals gave written informed consent.

## Data Availability

All data produced in the present study are available upon reasonable request to the authors

## Acknowledgement

The authors acknowledge the clinical staff of the Hepatopancreatobiliary Unit at Chris Hani Baragwanath Hospital, Johannesburg, South Africa for assistance with sample collection.

## Supplementary Material

**Supplementary table 1:** List of differentially expressed genes in tumours.

| Growth Factor array panel |  |  |  | Signalling pathway array panel |  |  |  |
| --- | --- | --- | --- | --- | --- | --- | --- |
| Upregulated |  | Downregulated |  | Upregulated |  | Downregulated |  |
| Gene | Fold regulation | Gene | Fold regulation | Gene | Fold regulation | Gene | Fold regulation |
| B2M | 190.39 | CSF2 | -7802.06 | HMOX1 | 261.74 | HEY2 | -5.66 |
| RPLP0 | 122.00 | HGDC | -6.84 | MCL1 | 162.02 | FABP1 | -4.50 |
| SPP1 | 73.73 | GDF10 | -3.86 | ADM | 91.77 | EPO | -3.71 |
| JAG1 | 60.65 | IL2 | -3.29 | B2M | 68.93 | TNF | -3.65 |
| ACTB | 41.30 | THPO | -3.10 | RPLP0 | 65.21 | HGDC | -3.57 |
| TYMP | 39.44 | TDGF1 | -3.04 | ATF4 | 45.73 | GATA3 | -3.49 |
| INHBA | 33.11 | CXCL1 | -2.63 | ACSL4 | 26.06 | OLR1 | -3.17 |
| IGF1 | 32.42 | BMP10 | -2.36 | FTH1 | 25.32 | IFNG | -2.96 |
| PTN | 28.32 | IL1B | -2.32 | BCL2 | 19.13 | WNT6 | -2.70 |
| GAPDH | 16.74 | NRG2 | -2.09 | CCL5 | 18.77 | NOTCH1 | -2.09 |
| PGF | 11.95 |  |  | ACTB | 17.98 |  |  |
| CSF1 | 10.30 |  |  | MMP7 | 16.61 |  |  |
| FGF7 | 9.42 |  |  | HES1 | 14.94 |  |  |
| PI | 8.31 |  |  | JAG1 | 14.57 |  |  |
| TGFB1 | 8.15 |  |  | SQSTM1 | 14.53 |  |  |
| PDGFC | 7.00 |  |  | CCND1 | 14.31 |  |  |
| FGF14 | 6.71 |  |  | MYC | 12.91 |  |  |
| BMP1 | 6.58 |  |  | CA9 | 12.89 |  |  |
| VEGFA | 6.34 |  |  | EGFR | 12.59 |  |  |
| FGF1 | 6.04 |  |  | HERPUD1 | 12.43 |  |  |
| MSTN | 5.70 |  |  | HEY1 | 11.93 |  |  |
| FGF2 | 5.67 |  |  | ID1 | 11.00 |  |  |
| GDF11 | 5.41 |  |  | TXN | 10.43 |  |  |
| NTF3 | 5.33 |  |  | NQO1 | 10.20 |  |  |
| IL18 | 4.90 |  |  | ICAM1 | 9.90 |  |  |
| FGF9 | 4.89 |  |  | CCND2 | 8.83 |  |  |
| TNNT1 | 4.59 |  |  | SERPINE1 | 8.08 |  |  |
| IL3 | 4.36 |  |  | CDKN1A | 8.06 |  |  |
| LIF | 4.28 |  |  | BAX | 7.96 |  |  |
| BMP5 | 4.12 |  |  | CDKN1B | 7.38 |  |  |
| BMP4 | 3.71 |  |  | SLC2A1 | 7.06 |  |  |
| CECR1 | 3.63 |  |  | GADD45B | 6.67 |  |  |
| BMP7 | 3.62 |  |  | GCLC | 6.08 |  |  |
| ERAP1 | 3.49 |  |  | PPARD | 5.87 |  |  |
| FGF13 | 3.48 |  |  | ACSL5 | 5.80 |  |  |
| IGF2 | 3.40 |  |  | VEGFA | 5.54 |  |  |
| NRG1 | 3.22 |  |  | FOSL1 | 5.40 |  |  |
| HBEGF | 2.92 |  |  | GAPDH | 5.32 |  |  |
| NODAL | 2.81 |  |  | ARNT | 5.10 |  |  |
| MDK | 2.81 |  |  | BCL2A1 | 5.07 |  |  |
| LTBP4 | 2.81 |  |  | EMP1 | 4.98 |  |  |
| IL12B | 2.81 |  |  | HEYL | 4.92 |  |  |
| FGF19 | 2.81 |  |  | LFNG | 4.25 |  |  |
| BMP2 | 2.81 |  |  | STAT1 | 4.11 |  |  |
| AMH | 2.81 |  |  | DAB2 | 4.08 |  |  |
| BMP3 | 2.49 |  |  | TNFSF10 | 3.94 |  |  |
| NRG3 | 2.44 |  |  | LRG1 | 3.91 |  |  |
| CSF3 | 2.35 |  |  | TXNRD1 | 3.44 |  |  |
|  |  |  |  | PCNA | 2.81 |  |  |
|  |  |  |  | BCL2L1 | 2.78 |  |  |
|  |  |  |  | CPT2 | 2.72 |  |  |
|  |  |  |  | WISP1 | 2.71 |  |  |
|  |  |  |  | LDHA | 2.70 |  |  |
|  |  |  |  | IRF1 | 2.68 |  |  |
|  |  |  |  | SORBS1 | 2.54 |  |  |
|  |  |  |  | GADD45A | 2.10 |  |  |

**Supplementary Fig. 1:**
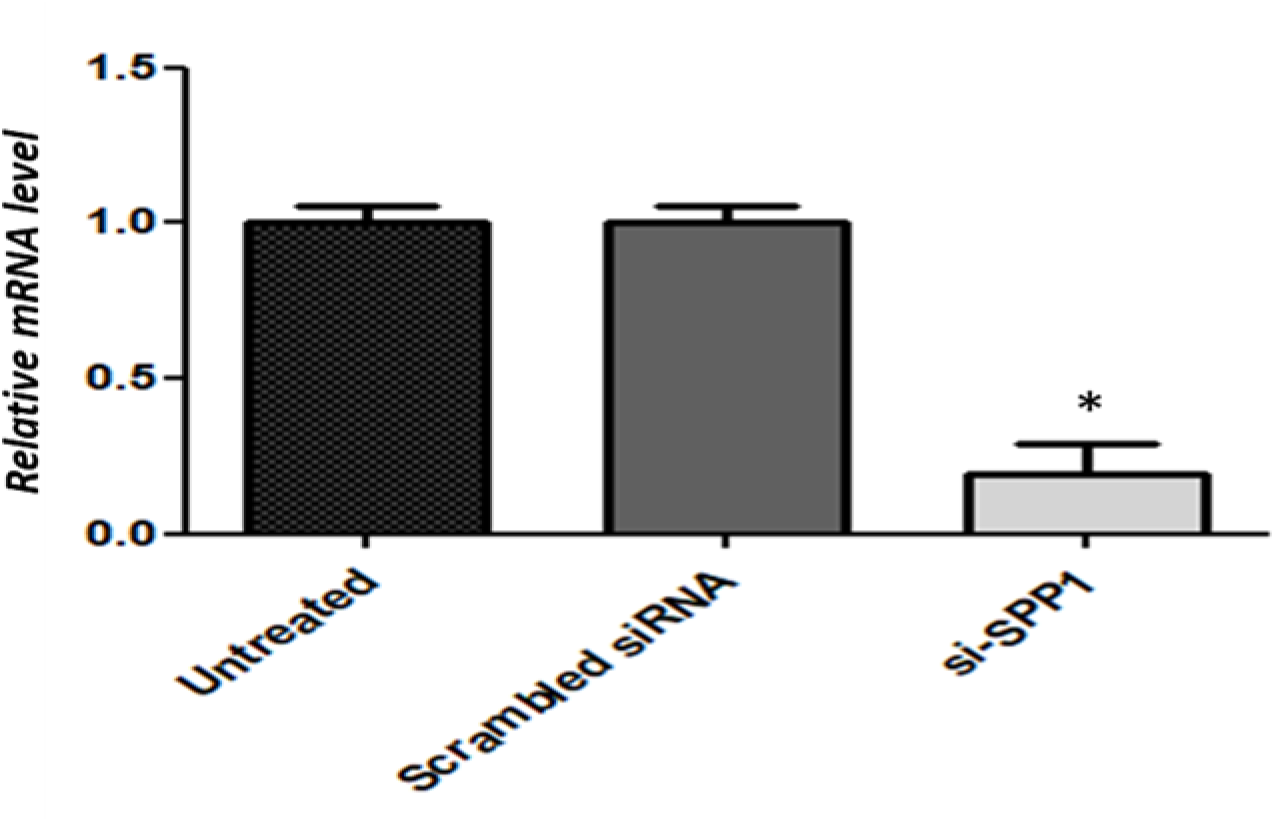
Validation of *SPP1* knockdown. *SPP1* knockdown of MIA PaCa-2 cells was performed for 48 h. Real-time PCR detected *SPP1* mRNA expression levels. β-Actin was used as a positive control for normalisation. As expected, the non-targeting control (NTC) siRNA did not significantly affect *SPP1* levels. *SPP1* levels were significantly decreased by 81% in the MIA PaCa-2 transfected cells. All values are given as the mean ± SD of three replicates. * denotes a p-value of less than 0.05.

**Supplementary Fig. 2:**
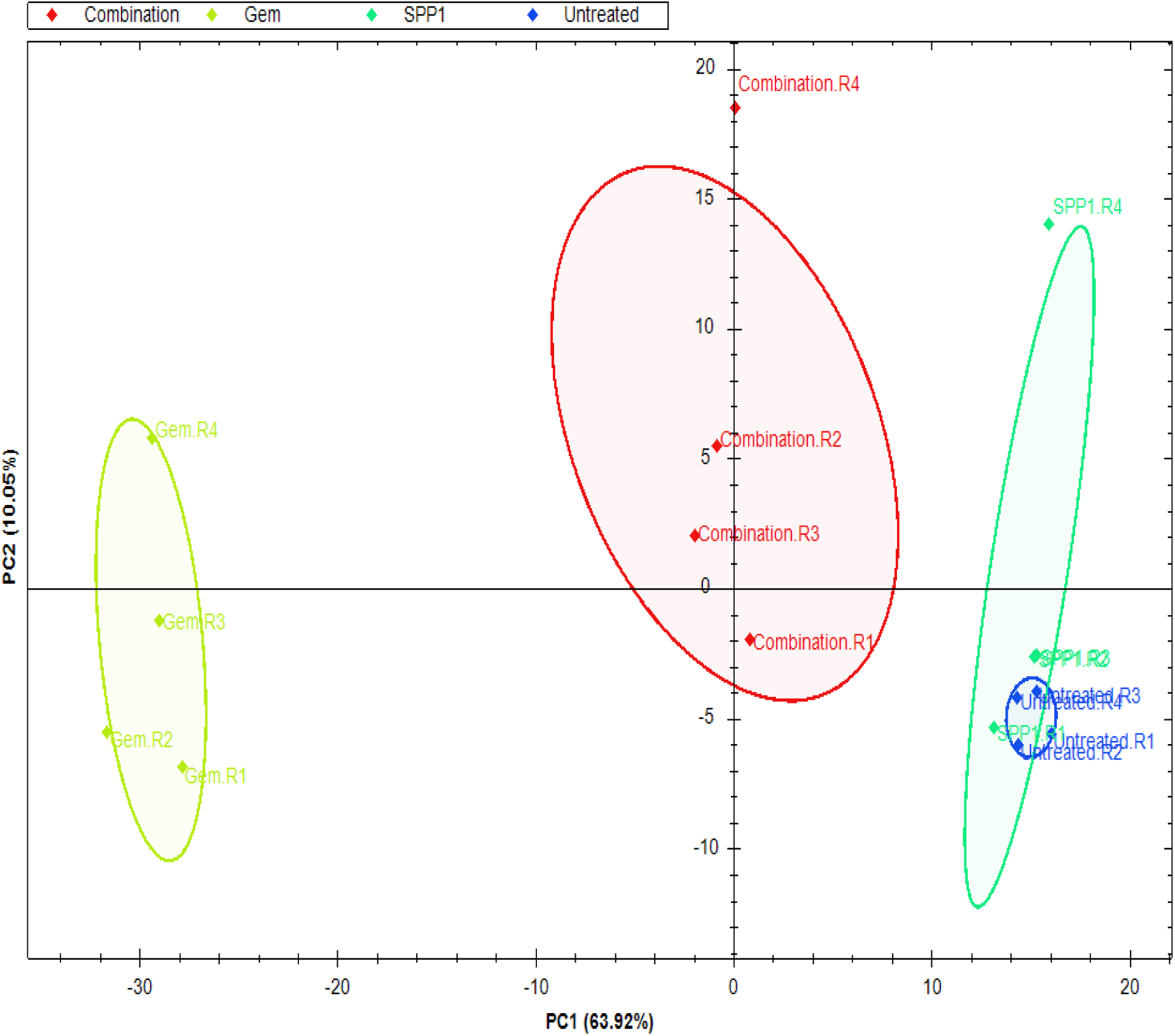
Principal component analysis (PCA analysis) conducted on the different treatment groups. PCA analysis indicates that the treatment groups are separated and well differentiated from each other. Although the *SPP1* knockdown and untreated groups overlap, the untreated group clusters tightly with PC2 having a greater influence over the *SPP1* knockdown group.

